# Modeling and Optimizing Deep Brain Stimulation to Enhance Gait in Parkinson’s Disease: Personalized Treatment with Neurophysiological Insights

**DOI:** 10.1101/2024.10.30.24316305

**Authors:** Hamid Fekri Azgomi, Kenneth H. Louie, Jessica E. Bath, Kara N. Presbrey, Jannine P. Balakid, Jacob H. Marks, Thomas A. Wozny, Nicholas B. Galifianakis, Marta San Luciano, Simon Little, Philip A. Starr, Doris D. Wang

## Abstract

Although high-frequency deep brain stimulation (DBS) is effective at relieving many motor symptoms of Parkinson’s disease (PD), its effects on gait can be variable and unpredictable. This is due to 1) a lack of standardized and robust metrics for gait assessment in PD patients, 2) the challenges of performing a thorough evaluation of all the stimulation parameters space that can alter gait, and 3) a lack of understanding for impacts of stimulation on the neurophysiological signatures of walking. In this study, our goal was to develop a data-driven approach to identify optimal, personalized DBS stimulation parameters to improve gait in PD patients and identify the neurophysiological signature of improved gait. Local field potentials from the globus pallidus and electrocorticography from the motor cortex of three PD patients were recorded using an implanted bidirectional neural stimulator during overground walking. A walking performance index (WPI) was developed to assess gait metrics with high reliability. DBS frequency, amplitude, and pulse width on the “clinically-optimized” stimulation contact were then systemically changed to study their impacts on gait metrics and underlying neural dynamics. We developed a Gaussian Process Regressor (GPR) model to map the relationship between DBS settings and the WPI. Using this model, we identified and validated personalized DBS settings that significantly improved gait metrics. Linear mixed models were employed to identify neural spectral features associated with enhanced walking performance. We demonstrated that improved walking performance was linked to the modulation of neural activity in specific frequency bands, with reduced beta band power in the pallidum and increased alpha band pallidal-motor cortex coherence synchronization during key moments of the gait cycle. Integrating WPI and GPR to optimize DBS parameters underscores the importance of developing and understanding personalized, data-driven interventions for gait improvement in PD.

## 1 Introduction

Gait disturbances are common symptoms of Parkinson’s Disease (PD) and can often manifest as decreased step length [1, 2], increased variability in step length and step time [3–5], and asymmetry between the two stepping legs [6]. Gait dysfunction reduces mobility, increases fall risk, and significantly impacts a patient’s quality of life [7–10]. While high-frequency deep brain stimulation (DBS) of the basal ganglia is highly effective at mitigating symptoms such as tremor, rigidity, and bradykinesia [11–16], its impact on gait is more variable and less predictable, with some reports showing improved gait [17–19], while others indicate no significant improvement or even worsening of gait [20–24]. These unpredictable effects of DBS settings, coupled with individual variability among patients, highlight the need for improved DBS treatments targeting advanced gait-related problems and understanding their impacts on the basal ganglia thalamocortical network neurophysiology [25].

One significant challenge in enhancing DBS outcomes for treating gait disorders is the lack of a standardized gait metric for clinicians to use during programming. Stride velocity, variability in step time and step length, and arm swing amplitude are among essential gait symptoms seen in PD patients [26–32]. However, focusing on a single metric may overshadow the comprehensive impact of DBS settings, leading to suboptimal outcomes. Currently, DBS programming is mainly conducted with the patient seated, focusing on the assessment of limb motor functions using the Unified Parkinson’s Disease Rating Scale (UPDRS). While some clinicians incorporate walking tests, these are often limited, non-standardized, and based primarily on subjective observation. Moreover, the UPDRS fails to capture many critical aspects of gait dysfunctions specific to PD, further complicating the evaluation and optimization of DBS settings for gait [33]. Thus, a systematic approach to assessing gait quality is necessary.

Even in cases where gait metrics are evaluated during DBS adjustments, identification of DBS stimulation parameters optimized for gait is difficult [17]. This is because the extensive parameter space (i.e., amplitude, frequency, and pulse width of the stimulation impulses at each contact) required to search for gait optimization would require an impractical amount of time for both patients and clinicians [34, 35]. Therefore, most clinicians primarily rely on empiric high-frequency stimulation during

DBS programming visits for clinical optimization. While high-frequency DBS is usually effective for tremor, rigidity, and bradykinesia, its efficacy for gait disorders is less consistent [36–38]. Lower frequency stimulation can improve gait kinematics, but significant uncertainties remain due to individual variations and inconsistent DBS effects [19, 22, 24, 39–44]. The variability in patient responses to different DBS stimulation parameters underscores the need for a more structured approach to systematically explore the full parameter space and identify settings that optimize gait functions [45–50].

The next challenge is the limited understanding of gait neurophysiology, which restricts our ability to fully understand the effects of DBS settings on modulating gait-related neural dynamics. Advances in sensing technologies now allow us to explore how changes in DBS settings influence the neural mechanisms driving motor functions [51–54]. By identifying neural biomarkers associated with gait improvements, we would gain 1) valuable insights into the circuits and structures involved in gait control, 2) understanding of how different stimulation parameters can result in similar improvements in gait, and 3) potentially leverage these biomarkers to guide DBS programming to target specific gait-related oscillations more efficiently to further enhance walking performance. Our previous study has shown that in PD patients without gait deficits, low-frequency local field potentials (LFPs) in the subthalamic nucleus (STN) and their synchrony with the primary motor cortex change cyclically based on the specific phase of the gait cycle. These dynamic oscillatory changes across the basal ganglia-cortical network may represent a physiologic signature of effective overground walking [55]. Despite these findings, the effects of DBS settings on neural signatures involved in gait are not well studied and may account for the variable effects of DBS on gait. Therefore, a deeper understanding of the relationship between DBS settings, underlying neural mechanisms, and resulting gait outcomes would further enhance our knowledge of the neurophysiological foundations of complex gait functions in PD.

Our study addresses these challenges by evaluating and modeling the effects of different DBS setting parameters on gait metrics and cortico-basal ganglia neurophysiology. We first developed a walking performance index (WPI) that integrates key gait kinematics to objectively assess and track gait performance across different DBS configurations. We then applied an individualized-Bayesian optimization method for modeling predicted walking performance based on the DBS settings for each subject, which efficiently delineates these relationships with limited trials. The data-driven model approach provided a robust tool to identify effective personalized DBS settings for gait improvement. Finally, by studying how DBS influences the pallidal and motor cortical network, we identified neurophysiological biomarkers associated with improved walking performance, which can further guide programming in the future (Figure 1). These findings significantly contribute to our understanding of DBS’s influence on gait disorders and support the development of personalized data-driven models of DBS parameter optimization that refine neuronal activity and enhance gait outcomes. This methodology could be adapted to address a variety of other symptoms within PD, potentially offering a framework for similar approaches in other circuit-based disorders.

**Fig. 1.**
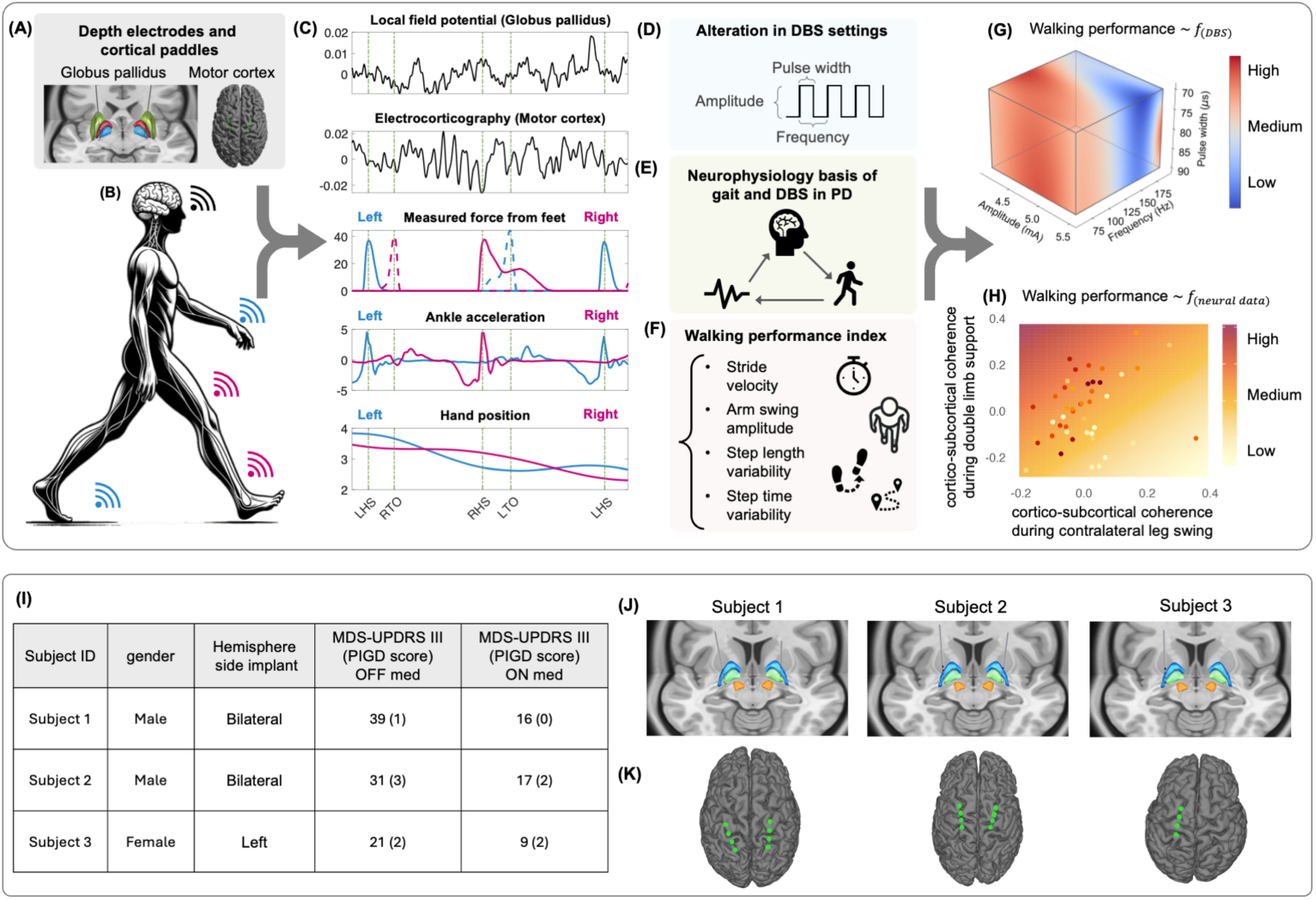
Workflow overview, patient characteristics, lead reconstruction, and electrode placement. Panel **(A)** illustrates the DBS depth electrodes implanted along with the cortical paddles placed to stimulate electrical impulses and capture neural activity. Panel **(B)** depicts a subject with Parkinson’s disease (PD) wearing a combination of sensors (i.e., Trigno system (Delsys) and MVN Analyze (Xsens)) to capture various gait kinematics. Panel **(C)** shows a sample of data collected during overground walking. The first two subpanels display neural data from the Globus pallidus and motor cortex. Next are the gait kinematics such as force sensors and ankle acceleration to capture gait events and spatiotemporal measurements to monitor body movements like arm swing amplitudes. Panel **(D)** outlines the components of stimulation parameters altered in our experiments. Panel **(E)** summarizes the goals of our analysis, which aim to understand the links between DBS settings, neurophysiological characteristics, and gait functions in PD. Panel **(F)** details the metrics employed to assess the impacts of DBS settings on walking performance, including variability in step length and step time, stride velocity, and arm swing amplitude. Panel **(G)** presents sample results from our data-driven approach, mapping the relationship between DBS setting parameters and walking performance, where deeper red colors indicate higher walking performance. Panel **(H)** exemplifies the outcome of our study in uncovering the neurophysiological bases of DBS settings associated with improved walking performance where the walking performance is a function of the cortical-subcortical coherence during different phases of the gait cycle. Panel **(I)** shows patient characteristics. MDS-UPDRS-III: Movement Disorders Society’s Unified Parkinson’s Disease Rating Scale; Part III: motor domain; PIGD: Posture Instability Gait Disorder (subscore from items 3.9: arising from the chair, 3.10: gait; 3.11: freezing, 3.12:postural instability, and 3.13 posture. Depth pallidal electrodes and cortical paddles over the motor cortex are displayed in Panels **(A)** and **(B)**, respectively.

## 2 Results

### Patient characteristics and electrode placement

Three participants with PD (1 female, 2 male) undergoing DBS implantation for motor fluctuations were recruited and implanted with a bidirectional investigational device (Summit RC+S, Medtronic). Two participants were implanted bilaterally, while one received a unilateral implant in the left hemisphere. All subjects had gait disturbances (Figure 1I). Gait disturbances in our subjects were assessed using the Movement Disorder Society’s UPDRS Part III (MDS-UPDRS-III), focusing on the motor symptoms associated with PD. Additionally, the posture instability and gait disorder (PIGD) sub-score provided a more specific evaluation of gait-related challenges, including difficulties with arising from a chair, walking, freezing episodes, postural instability, and overall posture [56]. All subjects were implanted with quadripolar DBS leads targeting the globus pallidus (GP) and quadripolar electrocorticography paddles placed in the subdural space over the motor cortical area. Figures 1J and 1K demonstrate the lead location reconstruction from both GP and motor cortex areas. In subject 1, the cortical paddle primarily records brain activity from the primary somatosensory (S1) area and M1 cortices. The cortical paddle spans the primary motor (M1) and premotor (PM) cortices in subjects 2 and 3. Each cortical paddle and the GP DBS lead in the same brain hemisphere were connected to a bidirectional neural stimulator device (summit RC+S, Medtronic Plc.). This device is used to deliver electrical impulses and can chronically stream high-resolution time-domain data [57].

### Walking performance index reveals changes in gait functions under different DBS stimulation parameters

To objectively evaluate overground walking metrics across all patients and assess their fluctuations in response to alterations in DBS settings, we developed a walking performance index (WPI), which is comprised of four kinematic parameters associated with common gait deviations in PD (Figures 2A and 2B): stride velocity (the average walking speed over a complete gait cycle), arm swing amplitudes, and variability in step length and step time. Higher WPI indicates quicker stride velocity, larger arm swing amplitudes, and lower variability in step length and step time. During each visit, we tested both clinically optimized settings (adjusted by each patient’s Movement Disorders neurologist) and several new DBS configurations by varying the stimulation frequency, amplitude, or pulse width within safe ranges. Patients performed overground walking in a 6-meter loop while their neural data and gait kinematics were continuously recorded. Each setting was evaluated over 200 steps, excluding turns, with turns evenly distributed between left and right. Gait metrics were calculated from full body inertial measurement unit (IMU) sensors that precisely capture gait kinematics.

**Fig. 2.**
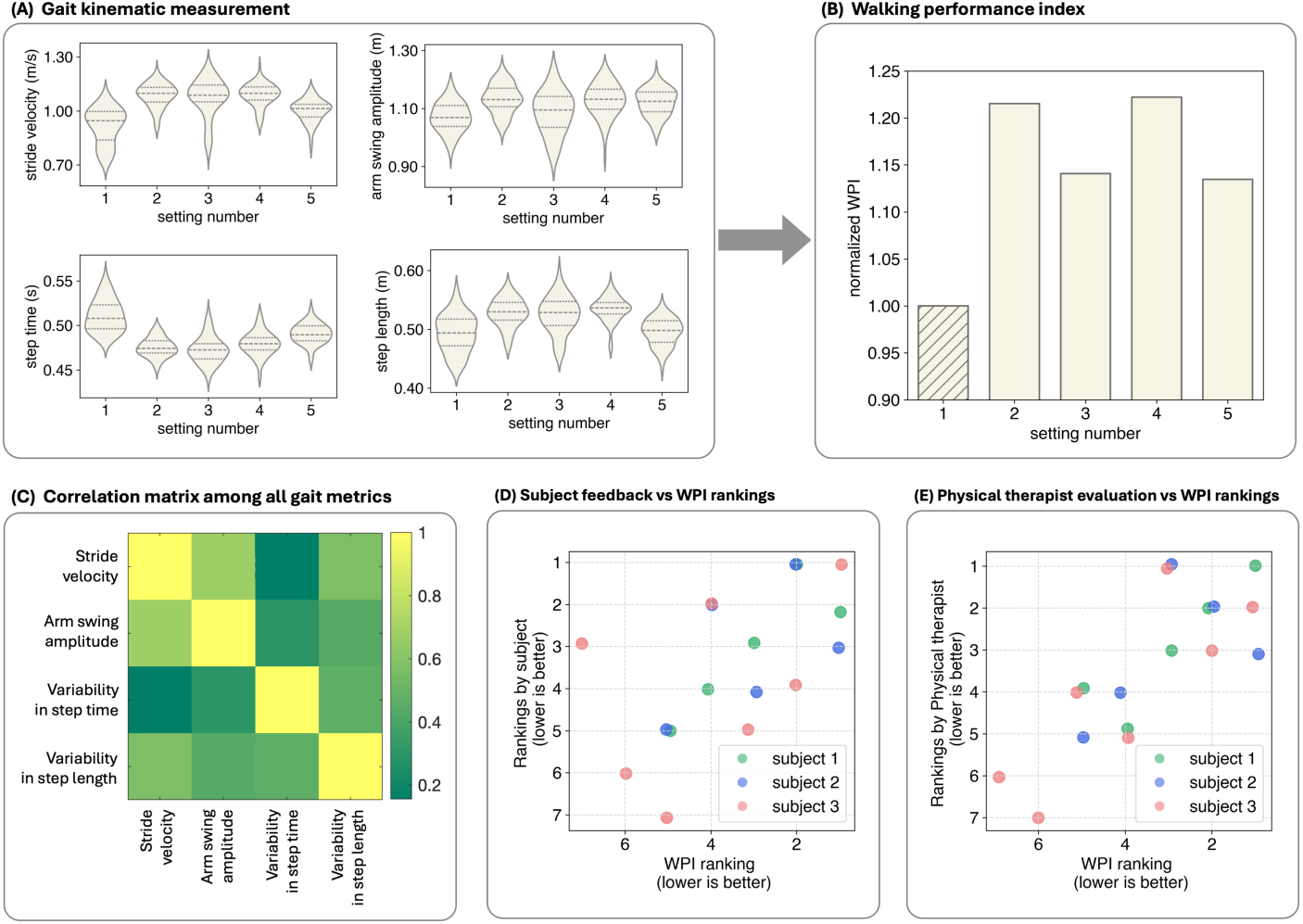
Walking performance development. Panel **(A)** shows an example of the variations in gait kinematics in response to different DBS settings. Panel **(B)** displays the corresponding fluctuations in the walking performance index (WPI). The column with the hatched pattern represents the clinically optimized DBS settings tested at each visit. Heatmap in panel **(C)** shows the pairwise Pearson correlation coefficients among all gait metrics and the WPI. The matrix visualizes the degree of linear relationships between variables, with color intensity representing the strength of the correlations: colors range from green (lower values) to yellow (higher values). The low off-diagonal values (green shades) indicate minimal multicollinearity among the metrics, confirming that each metric provides unique and independent information contributing to the WPI. Panels **(C)** and **(D)** show the correlation between the WPI rankings and feedback received from the subject, and evaluations from the physical therapist, respectively. Green, blue, and red dots represent data from subjects 1, 2, and 3, respectively. The x-axis displays the rankings based on WPI scores, while the y-axis shows the rankings provided by the patients. In both axes, lower numbers indicate higher rankings, with a rank of 1 representing the best performance or preference.

We assigned equal weights to each of the four gait parameters to ensure a balanced contribution from all variables and then normalized the WPI to that of their clinically optimized settings. Additionally, our analysis showed no signs of multicollinearity among the metrics, confirming that each metric provides unique information and contributes independently to the WPI (Figure 2C).

For each subject, we tested up to three amplitudes: clinical amplitude, 25-30% reduction from clinical and their higher limits (i.e., 4.1–5.5 mA for Subject 1, 2.8–5 mA for Subject 2, and 3.5–4.9 mA for Subject 3). In addition to the clinical frequency, we tested 60 Hz as well as a higher frequency setting (i.e., 60/190 Hz for Subject 1, 60/180 Hz for Subject 2, and 60/190 Hz for Subject 3). We also tested two pulse widths: clinical and their limits (i.e., 70/90 µs for Subject 1, 60/70 µs for Subject 2, and 60/80 µs for Subject 3). All these ranges were within the safety ranges defined by each patient’s neurologist. The details and range of DBS setting variables are presented in the bottom panels of Figures S1-S3.

As a result of changes in these DBS setting parameters, we observed significant modifications in subjects’ gait kinematics. Specifically, in Subject 1, stride velocity ranged from 0.5444 to 1.7217 m/s (Figure S1A). Arm swing amplitude ranged from 0.6571 to 1.5453 m (Figure S1B). Step time ranged from 0.3983 to 0.6075 s (Figure S1C). Step length ranged from 0.3511 to 0.8731 m (Figure S1D). The WPI varied from 0.4660 to 1.5597 (Figure S1E). In Subject 2, stride velocity ranged from 0.4738 to 1.2038 m/s (Figure S2A). Arm swing amplitude ranged from 0.3631 to 1.5121 m (Figure S2B). Step time ranged from 0.3105 to 0.7705 s (Figure S2C). Step length ranged from 0.2392 to 0.7897 m (Figure S2D). The WPI varied from 0.2656 to 1.3205 (Figure S2E). For Subject 3, stride velocity ranged from 0.6345 to 1.3132 m/s (Figure S3A). Arm swing amplitude, calculated as the average of both arms, ranged from 0.7215 to 1.5809 m (Figure S3B). Step time ranged from 0.4339 to 0.5689 s (Figure S3C). Step length ranged from 0.3605 to 0.6166 m (Figure S3D). These alterations in gait kinematics were accompanied by changes in the levels of WPI, which varied from 0.4352 to 1.4185 (Figure S3E).

Furthermore, participants were blinded to the changes in DBS settings during the trials. After each trial, their subjective feedback was collected to assess their perception of walking performance under different stimulation settings and compared to those of a blinded physical therapist. We show a good correlation between the WPI and the subject, as well as the physical therapist’s ranking of the settings (Figures 2D and 2E). This demonstrates the validity of the WPI to capture gait changes.

### Data-driven model identifies optimal DBS settings to improve gait in PD

To predict optimal DBS settings to enhance gait functions in PD, we employed a data-driven Gaussian process regressor (GPR) to map the relationship between the DBS settings (input) and the WPI (output) for each subject (see Figure 3). The GPR is particularly appropriate for this application because it can extrapolate data from a limited number of tested settings to predict outcomes across a broader parameter space. This approach allows for the efficient identification of optimal settings by leveraging the relationships between DBS parameters and gait performance, even when only a few settings have been experimentally tested. The GPR’s ability to model complex, nonlinear relationships and provide uncertainty estimates makes it beneficial for optimizing personalized DBS settings. We utilized the GPR after testing an average of 11 stimulation configurations to predict the next groups of settings to be tested. At each subsequent visit, we tested these model-predicted settings (best and worst) along with the clinically optimized setting. The resulting data were incorporated back into the GPR for further refinement.

**Fig. 3.**
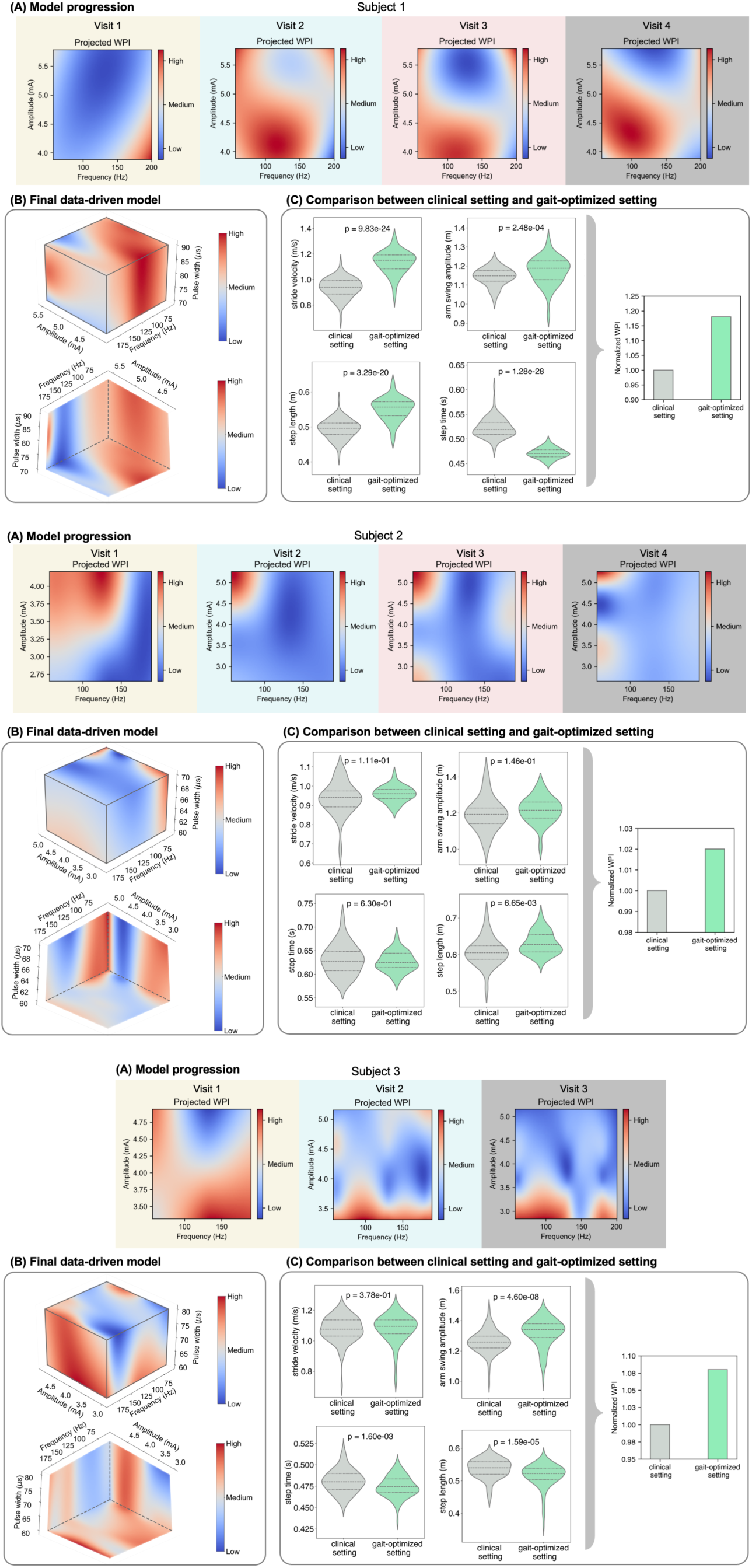
Data-Driven modeling and optimization of DBS settings. For each subject, panel **(A)** depicts the progression of the Gaussian Process Regression (GPR) model in 2-D planes, updated with data from each visit. Panel **(B)** showcases the final GPR model, mapping the relationship between DBS settings (amplitude, frequency, and pulse width) and WPI levels. The two perspectives illustrate the influence of amplitude, frequency, and pulse width on WPI levels, with red indicating higher walking performance and blue indicating lower WPI levels. Panel **(C)** compares WPI levels and associated gait kinematics under the clinical setting versus the gait-optimized setting derived from the GPR model.

The GPR model was able to predict and identify the optimal settings after testing several DBS configurations (i.e., 13 for subject 1, 20 for subject 2, and 11 for subject 3). Across all patients, the gait-optimized settings predicted by the model led to an overall improvement in WPI, with specific degrees of improvement varying by patient. Panels (A) in Figures 3 demonstrate the progression of the model between the DBS amplitude and frequency and WPI in 2D planes. In these plots, a deeper red color is associated with settings linked to higher walking performance levels. It illustrates how including the data from each visit would contribute to the changes in model dynamics, with the final model shown in Panels (B) of Figure 3. Each patient exhibited a unique gait-optimized setting. For Subject 1, the optimal setting of 5.1 mA, 60 Hz, and 90 µs showed a significant 18% improvement in WPI over the clinical setting of 5.5 mA, 150 Hz, and 90 µs (Figure 3B). Subject 2’s gait was optimized at 4.0 mA, 60 Hz, and 60 µs, compared to the clinical setting of 4.0 mA, 130 Hz, and 60 µs, leading to a modest 2% improvement in WPI (Figure 3B). For Subject 3, the model predicted that a setting of 4.2 mA, 180 Hz, and 80 µs would outperform the clinically optimized setting of 3.9 mA, 145 Hz, and 60 µs. Testing this prediction resulted in an 8% improvement in WPI (Figure 3B). Moreover, we observed a strong relationship between WPI scores and patient feedback rankings. The Spearman’s rank correlation coefficient was 0.62 (p = 0.008, Figure 2D).

There were also notable unique variations in individual gait parameters across the subjects (Panels (C) in Figure 3). Subject 1 exhibited the most pronounced improvements, with stride velocity increasing by 21.08% (Kruskal-Wallis, p < 0.0001) and arm swing amplitude by 2.71% (p = 0.0002). This subject also demonstrated a noticeable 46.83% reduction in step time variability and an 18.93% increase in step length variability (Figure 3C).

Subject 2 showed a 2.34% increase in stride velocity (p = 0.111) and a 1.98% increase in arm swing amplitude (p = 0.146, Figure 3C). Moreover, this subject experienced a substantial reduction in gait variability, with step time variability decreasing by 34.96% and step length variability by 19.96%. In Subject 3, stride velocity increased by 0.40% (p = 0.378), arm swing amplitude improved by 4.70% (p < 0.0001), and step time variability decreased by 11.22% (Figure 3C). However, step length variability increased by 22.34% in subject 1.

In Subject 1, after identifying and validating the gait-optimized DBS settings, we presented these parameters to their neurologist for clinical confirmation. After evaluating the optimized settings, the patient’s device was programmed with a new group with the gait-optimized settings. The patient was given the option to switch to this gait-optimized setting during longer walks and return to the clinical setting designed to manage their other symptoms as needed. An analysis of the device logs revealed that over a period of 64 days, the patient voluntarily used the gait-optimized setting for an average of 4 hours and 37 minutes per day. (Figure S4 in supplementary materials). These findings demonstrate the real-world applicability and efficacy of our proposed pipeline for identifying and implementing personalized DBS settings to improve gait functions, further validating its utility beyond controlled clinical environments.

### Identification of neural spectral changes across the gait cycle

To identify the neurophysiological basis for changes in these gait metrics, we analyzed pallidal LFP fluctuations, cortical activity, and pallidal-cortical coherence dynamics (i.e., magnitude-squared wavelet coherence) during each gait cycle as patients walked back and forth during the walking performance evaluation. Neural data streamed from the RC+S were time-synchronized to wearable devices that captured heel-strike and toe-off events. We first performed time-frequency and time-varying coherence analyses on each pallidal contact pair and the two cortical channels during all gait cycles, excluding turns, as standard gait cycle events do not occur during turning. To ensure the accuracy of our analysis, we meticulously removed all artifacts, including motion, electrocardiogram-related, and stimulation artifacts. Following artifact removal, we analyzed the spectral power within each frequency band and normalized the results across all tested DBS settings during each patient visit. Due to the presence of stimulation-induced sub-harmonic activity in some DBS settings, our analysis focused on the 1-30 Hz canonical frequency bands (i.e., delta (2-4 Hz), theta (4-8 Hz), alpha (8-12 Hz), and beta (12-30 Hz)) (see Methods section for additional details). Figure S5 illustrates an example of power spectral density and continuous wavelet transformation of the LFP signal in the GP region under 60 Hz and 145 Hz stimulation conditions for subject 3. By restricting our analysis to the 1-30 Hz frequency range, we ensured that stimulation-induced artifacts and their subharmonics were excluded from subsequent analyses (right panels of Figure S5A and Figure S5B).

The neurophysiological investigation of our study focused on identifying the average levels of signal power and coherence within distinct phases of the gait cycle and across canonical frequency bands. These features were extracted to capture the dynamic changes in neural activity and connectivity as patients moved through different stages of the gait cycle. An example of this analysis is presented in Panel (A) of Figure 4. Each gait cycle begins with the left heel strike (LHS) (purple line), followed by the right toe off (RTO) (orange line), right heel strike (RHS) (red line), and left toe off (LTO) (pink line), culminating in the next LHS. The phases of the gait cycle are defined as follows: the first double limb support period (DS1: LHS-RTO), contralateral leg swing (CLS: RTO-RHS), second double limb support period (DS2: RHS-LTO), and ipsilateral leg swing (ILS: LTO-LHS). As an example of characterizing the changes within canonical frequency bands across the gait cycle, we present the spectral power and coherence within the beta band (12-30 Hz) across all gait cycles (Figure 4B). Each row represents the average signal power or coherence within the beta band for a single gait cycle, sorted by stride time and initiated at LHS. This analysis revealed dynamic fluctuations in neural activity and coherence throughout the different phases of the gait cycle within each canonical frequency band (Figure 4C).

**Fig. 4.**
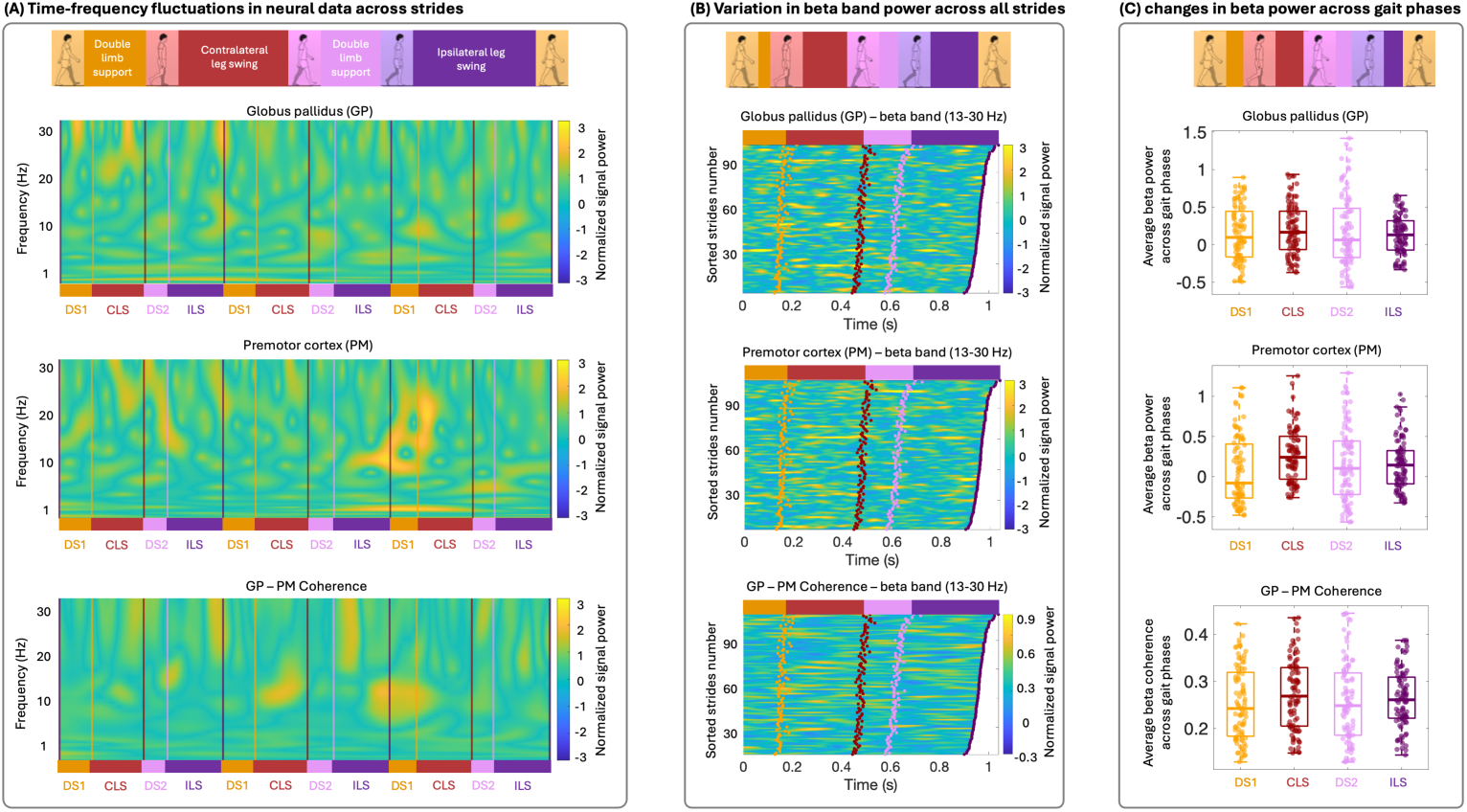
Neural data analysis and feature extraction. Panel **(A)** demonstrates a sample of continuous wavelet transformation of the neural data across three complete gait cycles from the GP (top), PM (middle), and GP-PM coherence (bottom) sub-panel from the left brain hemisphere, respectively. Each row shows the normalized values of a single frequency, initiated at the left heel strike (purple lines). Within each row, purple lines denote the left heel strike (LHS), orange lines denote the right toe-off (RTO), red lines denote the right heel strike (RHS), and pink lines denote the left toe-off (LTO) moments. Panel **(B)** demonstrates the average levels of signal power and coherence within the beta band (12-30 Hz) across all gait cycles, sorted by stride time and initiated at the LHS. Within each row, orange dots denote the RTO, red dots denote the RHS, and pink dots denote the LTO moments. All gait cycles conclude with the subsequent LHS (purple dots). Gait phases include DLS1 (double limb support 1) from LHS to RTO, CLS (contralateral leg swing) from RTO to RHS, DLS2 (double limb support 2) from RHS to LTO, and ILS (ipsilateral leg swing) from LTO to LHS. Boxplots in panel **(C)** represent average levels of the beta band during different phases of the gait cycle. Orange, red, pink, and purple colors, in turn, show the DS1, CLS, DS2, and ILS phases of the gait cycle.

### Personalized neural biomarkers associated with gait improvements

Investigating the relationship between neural spectral features —specifically, the average power levels during different phases of the gait cycle and within various canonical frequency bands— and walking performance, we conducted separate linear regression analyses for each neural feature. By analyzing each feature individually, we aimed to identify person-specific neural biomarkers associated with improvements in WPI. Table 1 highlights significant power bands (i.e., p < 0.05) within different gait phases observed for each individual, including data from both cortical and subcortical regions, as well as cortico-subcortical coherence. Additionally, coherence within each cortical region was assessed. These correlations were computed on aggregated data across all stimulation settings for each individual.

**Table 1.**
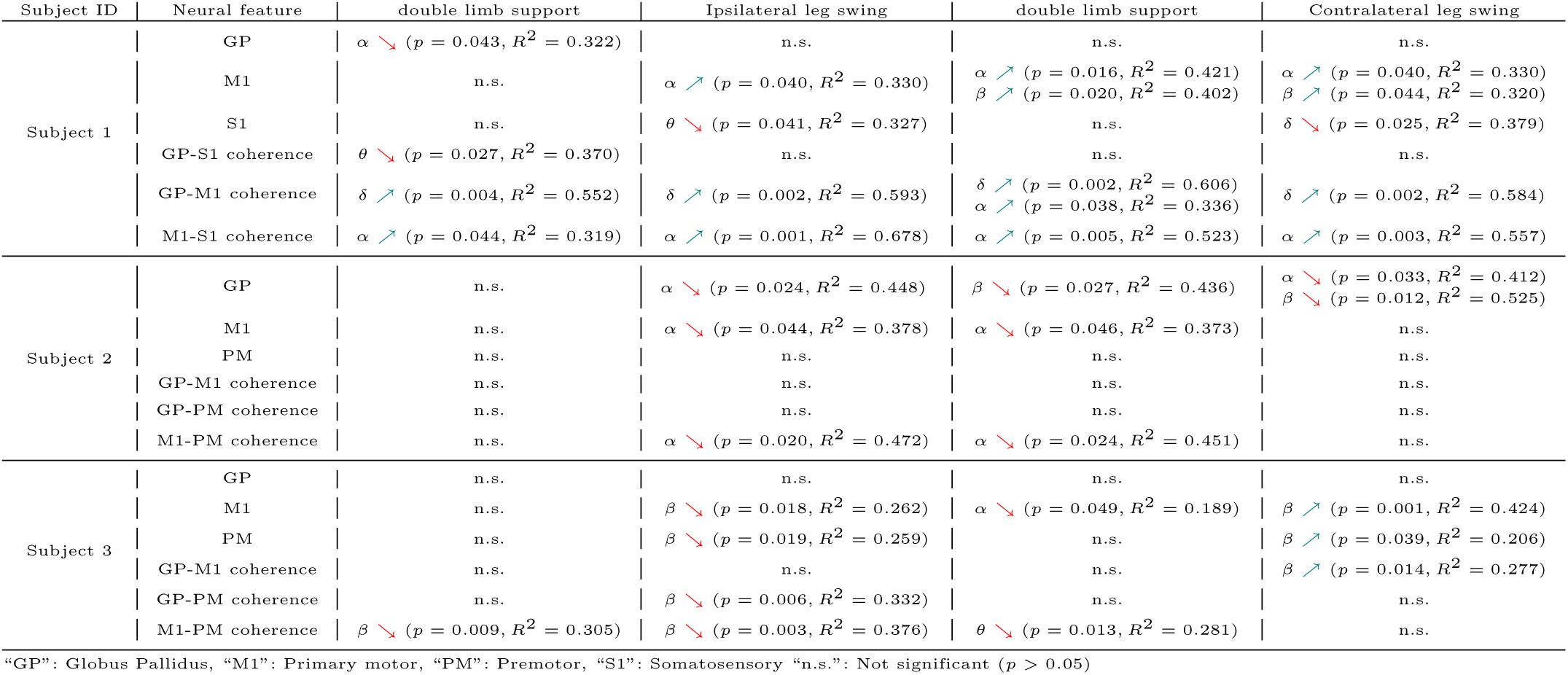
Person-specific neurophysiological characteristics.

Subject 1 exhibited a mix of positive and negative correlations between neural spectral features and walking performance. GP alpha-band power during double support was negatively correlated to walking performance (p = 0.043, R^2^ = 0.322), whereas M1 alpha power during contralateral leg stance (p = 0.040, R^2^ = 0.330) and beta power during contralateral leg swing (p = 0.044, R^2^ = 0.320) is positively correlated with WPI. Additionally, negative correlations were detected in the primary somatosensory cortex (S1) low-frequency power during both leg swing phases (ipsilateral: theta: p = 0.041, R^2^ = 0.327; contralateral: delta: p = 0.025, R^2^ = 0.379). Coherence analyses revealed both negative and positive correlations between various regions and frequency bands. Notably, significant positive correlations were found between GP-M1 delta-band coherence across all gait phases (p < 0.005, R^2^ < 0.5) as well as M1-S1 alpha-band (p < 0.05, R^2^ > 0.3). Detailed correlations are listed in Table 1.

In Subject 2, significant negative correlations were primarily observed between GP alpha-power and walking performance during leg swing (ipsilateral: p = 0.024, R^2^ = 0.448; contralateral: p = 0.033, R^2^ = 0.412) and GP beta power during double support (p = 0.027, R^2^ = 0.436). During ipsilateral leg swing and double support periods, both M1 alpha power (p < 0.05, R^2^ > 0.35) and M1-PM alpha band coherence (p < 0.05, R^2^ > 0.45) were negatively correlated with the WPI. These results indicate that reductions in alpha and beta power in GP and M1 are associated with improved walking performance for this subject. Refer to Table 1 for detailed statistical values. For Subject 3, significant correlations were observed between neural spectral features in motor cortical regions and walking performance. Both negative and positive correlations were found in the primary motor cortex (M1) and premotor (PM) areas during specific gait phases and frequency bands. For example, a negative correlation between M1 beta-band power during the ipsilateral leg swing phase and walking performance was identified (p = 0.018, R^2^ = 0.262), while a positive correlation was observed during the contralateral leg swing phase (p = 0.001, R^2^ = 0.424). Similar patterns were seen in pallidal-cortical coherence, with significant correlations in globus pallidus GP-M1 and GP-PM coherence varying by gait phase and frequency band. Notably, a negative correlation was found between beta coherence within the M1-PM regions during the ipsilateral leg swing phase and walking performance (p = 0.003, R^2^ = 0.376). These findings suggest phase- and frequency-specific neural patterns associated with walking performance in this individual. Detailed results are provided in Table 1.

Across all subjects, the significant correlations varied by brain region, frequency band, and gait phase, underscoring the individuality of neural mechanisms underlying gait improvements. While some common trends were observed—such as the involvement of alpha and beta bands in motor cortical regions—the specific patterns differed among individuals. A comprehensive analysis of all neural features, including those that did not reach statistical significance, is presented in the supplementary materials (Figure S6). This broader view provides additional insights into the relationships between neural spectral features and walking performance across all individuals.

### Shared neural spectral features correlate with walking performance across subjects

To identify shared spectral features associated with improvements in gait performance across individuals, we used a linear mixed model to evaluate neural biomarkers of walking performance. Given the exploratory nature of our analysis, we focused on assessing correlations between individual neural features and walking performance, rather than constructing a comprehensive model incorporating all potential features simultaneously. By analyzing each feature separately, we aimed to identify consistent neural biomarkers across all participants that correlate with walking performance. This model included fixed effects for each neural feature and random effects for each individual. We extracted and averaged spectral power across all gait cycle epochs and assessed the correlation with the average levels of walking performance across all participants, covering multiple visits and different DBS settings. We observed a significant negative correlation between the beta band LFP signal power in the pallidal region and the increase in overall walking performance (Figure 5A). This significant correlation was primarily seen in the beta band power during the ipsilateral leg swing and double limb support following the contralateral leg swing. Specifically, in the double limb support phase, beta band power was found to be associated with walking performance, with higher performance associated with lower beta band power (Estimate = -0.184, SE = 0.0855, t(42) = 2.15, p = 0.037, *x*^2^ ≈ 4.37, p = 0.036). Similarly, during the ipsilateral leg swing phase, beta band power was found to be associated with walking performance (Estimate = -0.183, SE = 0.0859, t(42) = 2.13, p = 0.039, *x*^2^ ≈ 4.29, p = 0.038). In both phases, the inclusion of beta band power significantly improved the model fit over the null model. These findings suggest that a reduction in beta band power in the pallidal region during the contralateral leg stance phase is a critical neural marker for improved gait outcomes.

**Fig. 5.**
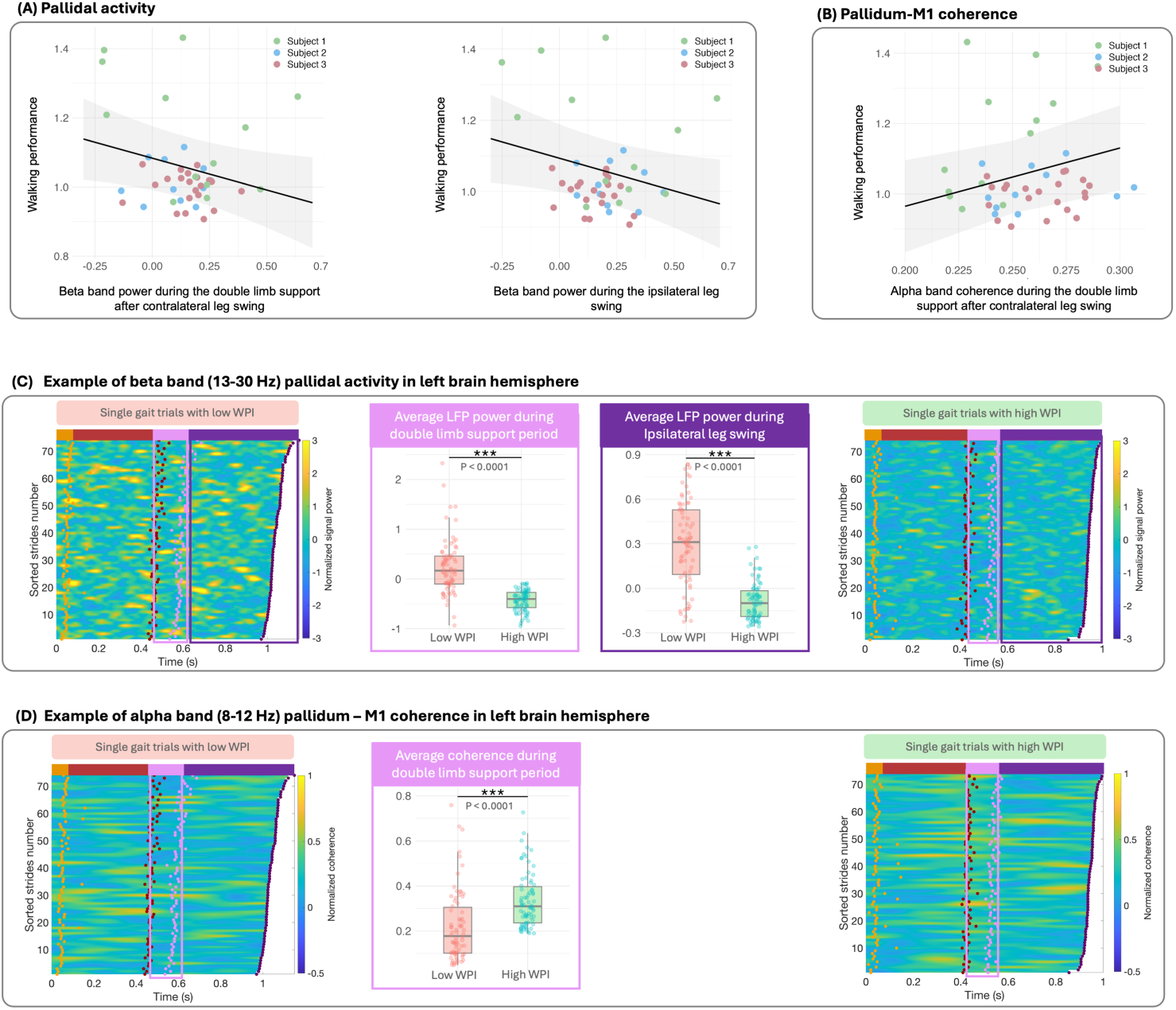
Correlation analysis of neural signal characteristics and walking performance. Panel **(A)** presents the relationship between changes in LFP signal power in the pallidal area and walking performance. Left and right sub-panels, in turn, show the relationship between the walking performance and the beta band power during the double limb support period after the contralateral leg swing and the ipsilateral leg swing, respectively. Panel **(B)** illustrates the relationship between the alpha band power of cortical-subcortical coherence (i.e., pallidum and M1) during the double limb support period after the contralateral leg swing and walking performance. Panels **(C)** and **(D)** exemplify the comparison of the neural signal variations from the left brain hemisphere between the sessions with low and high WPI across all single gait cycles. Panel **(C)** illustrates the beta band power of LFP signal power in the pallidal area, while panel **(D)** shows the alpha band power of pallidum–M1 coherence. In each panel, heatmaps on the left and right correspond to the lowest and highest walking performance, respectively. Each row represents a single gait trial, sorted by stride time and initiated at the left heel strike. Orange dots denote the right toe-off, red dots denote the right heel strike, and purple dots denote the left toe-off. All gait cycles conclude with the subsequent left heel strike. Middle sub-panels compare signal power during the specific gait phases of interest (i.e., double limb support period and ipsilateral leg swing in panel **(C)**, and double limb support period in panel **(D)**).

Similarly, we performed a thorough analysis to assess how the interaction between the pallidum and primary motor (M1) cortex could be associated with improvements in walking performance. We analyzed the averaged magnitude-squared coherence value between the pallidum and primary motor cortex (M1) for each brain hemisphere during different gait phases. We observed a positive correlation between pallidum-M1 coherence and increased walking performance (Figure 5B). This significant correlation was primarily observed in alpha band power during double limb support following the contralateral leg swing. Specifically, higher alpha band power in the interaction between the pallidum and M1 during double limb support is linked to higher walking performance (Estimate = 1.66, SE = 0.72, t(43.03) = 2.32, p = 0.025, *x*^2^ ≈ 4.37), and its inclusion significantly improved the model fit over the null model, as indicated by the likelihood ratio test (p = 0.029, *x*^2^ ≈ 4.79). These findings suggest that elevation in cortico-pallidal alpha band coherence serves as a promising neural marker for improved gait dynamics.

To further illustrate these neurophysiological characteristics and their relationship with walking performance, we compared neural activity between sessions with the lowest and highest WPI levels in subject 1 (Figures 5C and 5D). These figures highlight key differences in beta-band power in the pallidal area as well as alpha-band pallidum-M1 coherence in the left hemisphere. The heatmaps on the left side of Figures 5C and 5D display the neural activity associated with the lowest walking performance, while those on the right correspond to the highest performance. The middle sub-panels provide a more focused comparison of signal power during key phases: double limb support after the contralateral leg swing and ipsilateral leg swing in Figure 5C, and double limb support after the contralateral leg swing in Figure 5D, contrasting sessions with the lowest WPI (represented by red boxplots) and the highest WPI (represented by green boxplots). These visualizations underscore the distinct neural dynamics that differentiate optimal gait performance from less effective patterns.

## 3 Discussion

We developed a data-driven pipeline to identify optimized DBS setting parameters for enhancing gait in individuals with PD. Our research is one of the first to systematically identify optimized DBS settings (using amplitude, frequency, and pulse width) with a personalized data-driven approach targeting several spatiotemporal aspects of gait. The proposed model maps the relationship between DBS settings and walking performance across a wide range of parameter values, allowing us to predict and identify optimal stimulation configurations beyond the discrete settings initially tested, thereby significantly enhancing walking performance. Additionally, we identified neural biomarkers associated with improved walking performance, revealing both consistent characteristics across patients and features specific to each individual. The proposed pipeline links DBS configurations, underlying neural connectivity, and gait kinematics. Our results have several implications which are discussed below.

In this study, we developed an objective walking performance index (WPI) based on kinematic measures to accurately assess the effectiveness of our approach. The WPI represents core aspects of gait often impaired in PD, which can lead to significant functional limitations and increased fall risk [2, 4, 26–30, 42, 58]. Stride velocity indicates overall mobility, with reduced speed being a common symptom of PD [29, 32]. Arm swing amplitude reflects the coordinated movement necessary for balance [30], while variability in step length and time provides insight into gait consistency, with greater variability often linked to instability and fall risk [26]. By combining these metrics, the WPI offers a more comprehensive assessment of gait, addressing multiple dimensions of motor function affected by PD. The use of equal weighting in the WPI ensures a balanced contribution from all gait parameters, preventing any single metric from disproportionately influencing the overall score. Our findings support this approach, as each metric contributed unique information without multicollinearity issues. It is also worth mentioning that alternative weighting schemes could affect the sensitivity of the WPI to specific aspects of gait dysfunction. Future research might explore differential weighting based on the relative impact of each parameter on functional outcomes or patient quality of life.

The WPI’s broad evaluation of gait characteristics allows for a more robust assessment of DBS settings and their effect on walking. Our results confirmed that changes in DBS settings were effectively captured by the WPI, aligning with patient and clinician evaluations during each visit. This validation supports that the WPI is an effective metric for assessing and targeting gait improvements in PD. The WPI provides a quantifiable measure to track improvements and identify optimal DBS settings for each patient, potentially enhancing the precision of DBS programming. Future directions include developing automated systems for real-time gait analysis and integrating WPI with DBS programming software. Technologies such as gait mats, wearable sensors, and advanced motion capture systems could enable continuous and precise monitoring of gait, allowing for more accurate DBS adjustments. Further studies should explore the WPI’s broader application across different populations and its potential to improve clinical outcomes in PD and other movement disorders.

Optimizing DBS settings to enhance gait in individuals with PD is challenging due to the vast parameter space and the time-intensive process required during patient visits. While programming DBS for motor symptoms like bradykinesia and tremor typically yields quick results, gait disturbances are more complex and may take longer to respond, adding to the complexity of clinic visits. Moreover, individual responses to DBS therapy vary significantly, necessitating a systematic, personalized approach to identify optimized settings. We hypothesized that the extensive DBS parameter space, combined with the complex nature of gait and individual variability, requires a personalized, data-driven approach. Unlike methods that compare discrete stimulation parameters, we aimed to establish a continuous map between DBS settings and walking performance. We employed a Gaussian process regressor (GPR) to model these dynamics, using data from each visit with DBS settings as input and the WPI as output. The GPR model, with its non-parametric nature, effectively captured the complex relationship between DBS settings and WPI by updating its predictions based on continuous variations in DBS settings.

Our model incorporated several stimulation parameters—amplitude, frequency, and pulse width—as inputs, enabling us to understand the impact of each parameter in relation to the others on walking performance. One key advantage of this approach is its ability to explore the entire DBS parameter space (i.e., amplitude, frequency, and pulse width) within safety ranges defined by neurologists, potentially deriving optimized settings beyond the parameters tested. Contact selection was kept constant and determined separately based on each patient’s movement disorder neurologist’s expertise, rather than being optimized by the model. Testing the identified settings further validated the model’s efficacy, resulting in significant improvements in walking performance. This approach could enhance the DBS programming process by better tracking the effects of changes in stimulation parameters on each spatiotemporal gait metric [59]. While our current pipeline still requires several visits by the patient, with the identification of key neurophysiological signatures associated with enhanced gait functions, we envision using advanced machine learning models in the future that can potentially predict stimulation parameters that boost these oscillations without the need for extensive tests. To further enhance the model and expedite the optimization process, reinforcement learning algorithms could dynamically adjust DBS settings in real-time based on continuous gait performance feedback, while deep neural networks could capture complex, non-linear relationships between DBS parameters, neurophysiological dynamics, and gait performance, potentially improving prediction accuracy. Transfer learning could also utilize pre-trained models on similar datasets, reducing the data needed for training and accelerating optimization for new patients.

An important finding of our work is that although patients’ gait-optimized settings are varied, there are consistent and convergent neural dynamics identified in these settings that are shared on a group level. Specifically, lower levels of pallidum beta-band activity during the contralateral stance phase were associated with improved walking performance. Previous studies have shown that beta power in basal ganglia regions correlates with PD off-symptom severity and that its suppression through dopaminergic therapy or DBS improves UPDRS scores [60–66]. Specifically during gait, reduced beta-band activity in the GPi is linked to improved performance [52], and STN DBS can reduce high beta frequency power and bilateral oscillatory connectivity during gait [67, 68]. This reduction in beta activity may reflect a general mechanism of release from motor inhibition, facilitating smoother, more coordinated, and faster movements [69]. Additionally, increased alpha band coherence between the pallidum and the primary motor cortex (M1) during the double limb support phase following the contralateral leg swing was identified as a potential neural biomarker for enhanced walking performance. Alpha frequency modulation during gait phases has been described in the pedunculopontine nucleus (PPN) and cortex in PD patients, and is associated with increased regularity of stepping [70, 71]. We expand on these findings and identify a potential network oscillatory biomarker of gait improvement in response to DBS stimulation. Our discovery of increased alpha coherence between the pallidum and motor cortex at the beginning of the contralateral leg stance (double support period), followed by pallidal beta desynchronization during the contralateral leg stance phase, may represent a network mechanism to improve stepping regularity and mechanics. Our findings deepen the understanding of the neural mechanisms underlying DBS-enhanced gait performance and emphasize the need to identify individualized DBS settings to enhance these patterns.

Our study highlights the importance of oscillatory activity during gait functions and the potential of these neural biomarkers to differentiate between normal and impaired gait for a more effective therapy. Unlike established approaches that track biomarkers associated with other PD symptoms—such as beta suppression for akinesia and rigidity or gamma-band activity for dyskinesia—there is a notable lack of established biomarkers specifically linked to gait dysfunction in PD. By uncovering network oscillatory activities associated with gait enhancement, our findings provide valuable insights that could facilitate the programming process and open avenues for developing adaptive, closed-loop DBS systems. These systems require a comprehensive understanding of the impacts of DBS on pathological neural oscillations and behavioral measurements. The insights gained from our study could enable the future design of closed-loop DBS systems that monitor neural oscillatory activity in real-time and adjust stimulation parameters accordingly to modulate pathological oscillations, ultimately leading to improvements in walking performance. In such human-in-the-loop systems, neural oscillations would serve as feedback signals in a decision-making platform where DBS parameters are adjusted based on real-time tracking of neural activity. Advancing our understanding of the links between DBS settings, underlying neural oscillations, and gait outcomes could pave the way for control system–based methodologies that adjust stimulation parameters in real-time. Such developments hold significant potential not only for improving gait in PD but also for addressing motor symptoms in a wide range of neurodegenerative diseases.

Our study has limitations. The data-driven model’s performance was validated with a small number of participants, and future studies with larger populations are needed to confirm its efficacy in identifying optimized DBS settings with fewer trials. We did not impose constraints on total energy delivery, which could be a future optimization target. Additionally, we relied on neurologists’ selections for GP stimulation contacts without exploring their effects in detail. Our analysis focused mainly on straight walking, suggesting that future models should incorporate factors like turning and gait initiation to improve treatment outcomes. Our exploratory analysis identified neural biomarkers associated with gait improvements at both the group and individual-specific levels. However, we recognize that the limited sample size and the investigative nature of this study may affect the generalizability of our findings. Future studies with larger cohorts and more extensive data collection are warranted to validate these neural biomarkers and to further interpret the neural mechanisms underlying gait improvements in response to DBS.

## 4 Conclusion

Our study systematically modeled the relationship between DBS setting parameters, underlying neural dynamics, and gait functions, demonstrating the efficacy of datadriven techniques in optimizing patient-specific DBS settings. By integrating key gait kinematics into our performance assessment, we developed a robust tool for enhancing motor function through personalized DBS interventions. Our data-driven model effectively identified optimal DBS settings tailored to each patient, resulting in significant improvements in gait performance. The neural biomarkers we identified could be used to assess the impact of DBS settings on gait functions, providing insights for further optimization of stimulation parameters. This approach not only deepens our understanding of how DBS settings modulate gait but also highlights the potential for personalized treatment strategies applicable across a wide range of neurodegenerative diseases. The identified neural biomarkers have the potential to be utilized in electrophysiology-based optimization of DBS settings targeting gait functions. The methodologies explored in this research underscore the importance of addressing individual patient needs to achieve more effective DBS therapies.

## 5 Methods

### Subjects’ recruitment, DBS surgery, and electrode localization

Three participants with Parkinson’s disease (PD) and gait dysfunction (1 female, 2 male, age range: 48-66 years, disease duration: 3-13 years) were recruited and implanted with Medtronic Summit RC+S devices (two subjects received bilateral and one received unilateral), with subdural paddle electrodes over the motor cortex and deep brain stimulation (DBS) electrodes in the globus pallidus (Figures 1J and 1K). Participants were enrolled in a clinical trial (ClinicalTrials.gov ID: NCT-03582891) at the University of California San Francisco (UCSF), which was approved by the UCSF Institutional Review Board (IRB) under the approval number 20-32847. The leads were attached to a research-grade, sensing-capable implantable pulse generator (Medtronic Summit RC+S model B35300R), which was positioned in a compartment over the pectoralis muscle on each side. Precise electrode localization was achieved through established image analysis pipelines for both depth and cortical electrodes. Post-implantation high-resolution CT images were briefly coregistered to preoperative T1-weighted 3T MRI using a rigid, linear affine transformation. The accuracy of coregistration was verified through visual inspection and, when necessary, refined using an additional brain shift correction routine to align subcortical anatomy. Electrode artifacts were then identified on CT and matched to known electrode geometry. Additionally, cortical electrodes were projected onto the MRI-rendered pial surface. More details regarding the surgical implantation and lead reconstruction can be found in [57, 72–75].

### Experiment procedures and data collection

For each subject, the DBS setting configuration was optimized before initiating the tasks. This optimization (their “clinical setting”) was completed by each patient’s Movement Disorders neurologist over a range of 1.5 – 3 months. All subjects were receiving their typical dose of Parkinsonian medication throughout data collection and follow-ups. In all participants, local field potentials (LFPs) were recorded from the globus pallidus (GP). Electrocorticography (ECoG) recordings were recorded from cortical sites were carried out using pairs of contacts: 9 and 8 for either the primary motor (M1) or somatosensory cortices (S1), and 10 and 11 for either the premotor cortex (PM) or M1, as determined by imaging-based reconstruction (Figures 1J and 1K). Neural data were acquired at a sampling rate of 500 Hz. Moreover, the RC+S system’s built-in accelerometer, which synchronizes measurements with external sensors, collected accelerometry data at 64 Hz. We extracted and analyzed all the data using open-source code available in https://github.com/openmind-consortium/Analysis-rcs-data [76].

During clinic visits, patients’ DBS settings were altered within safety ranges to examine their impacts on modulating gait functions. In response to each set of DBS settings, participants performed overground walking in a loop of approximately 6 meters with continuous streaming of their neural data and gait kinematics. Each setting was tested for 200 (non-turning) steps, with half the turns to the left and half to the right. Before each walking trial, the research team systematically changed the stimulation settings. These adjustments encompassed changes in stimulation amplitude, frequency, and pulse width. A minimum interval of 15 minutes was observed between each walking trial to allow a proper washout period. Following the completion of the overground walking tasks, subjects were instructed to sit and rest for three minutes. After this initial rest period, a member of the research team collected patient feedback using standardized questions covering the presence of any potential side effects, perceived effort with walking, gait symmetry, perceived balance and/or stability, and the overall feel of the setting compared to everyday function. Upon receiving their feedback, participants were asked to sit and rest as they received new sets of stimulation parameters. All trials were conducted in a randomized order with examiner and patient blinding.

We used two types of wireless technology to capture gait kinematics: Trigno system (Delsys) and MVN Analyze (Xsens). Delsys included two Avanti force-sensitive resistor (FSR) adapters, two Avanti goniometer adapters, and two Trigno surface electromyography (EMG) sensors with built-in accelerometers. EMG sensors were placed on the lower legs (bilateral soleus and tibialis anterior muscles) for precise muscle activity measurement. Each FSR adapter connects to four FSRs (model DC: F01, Delsys) positioned under the heel (calcaneus), big toe (hallux), base of the big toe (first metatarsal), and base of the pinky toe (fifth metatarsal). A digital goniometer (SG110/A) was placed next to the ankle bone (lateral malleolus) on each side to measure continuous ankle joint angles. Xsens uses 15 body-worn sensors to detect position and movement for full-body motion tracking [77]. Subjects were also video recorded with multiple camera views to facilitate additional inspection and analyses.

### Integrating gait kinematics with walking performance index

This study focuses on straight-line overground walking; hence, turns were excluded from the analysis. Gait kinematics, including step length and step time duration, are derived utilizing Delsys and Xsens recordings. The initial detection of gait events necessary for our analysis is conducted using FSR sensors. However, in instances where gait peculiarities, such as toe walking, result in suboptimal signal strength, alternative methods (e.g., goniometer signal) were employed to ensure accurate identification of gait events. All gait events underwent visual inspection, with any inaccuracies manually adjusted. To analyze how walking performance is influenced by changes in DBS settings, we developed a walking performance index (WPI) to capture various gait kinematics, including stride velocity, step length variability, step time variability, and arm swing amplitude. These metrics were selected based on their importance in the literature on PD gait [26–32]. During each visit, we normalized the WPI relative to the baseline levels observed in the clinical setting.

### Data-driven model development and identification of gait-optimized DBS setting

Leveraging data-driven methodologies, we modeled the relationship between distinct DBS settings as the input and the WPI as the output. Employing the Gaussian process regression (GPR) method, we created personalized maps detailing how DBS settings influence WPI for each participant, highlighting which stimulation settings could enhance WPI. GPR stands out as a flexible, non-parametric Bayesian approach, adept at capturing complex relationships between variables without presupposing the form of the underlying function [78]. This method excels in scenarios where the relationship’s exact nature is unknown or too complex to model with traditional parametric techniques. By integrating a potentially infinite number of parameters, GPR allows the data to dictate the complexity of the model, adapting to the nuances of each individual’s response to DBS adjustments. Furthermore, GPR’s utility extends beyond passive analysis, facilitating active learning by optimizing input selection to maximize desired outcomes, making it an invaluable tool in the iterative process of identifying optimal DBS settings for enhancing gait performance. This approach aligns with previous studies across various domains, demonstrating the versatility and power of GPR in both understanding and optimizing human behavior and physiological responses. In our implementation, we adopt the Matern kernel as the covariance function [79], a choice driven by its flexibility and capability to capture the roughness of the functional relationship between DBS settings and WPI. The Matern kernel is defined as:

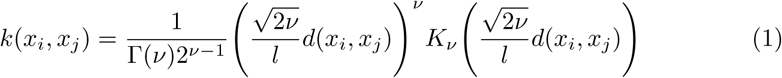

where d(., .) is the Euclidean distance, *K_v_* (.) is a modified Bessel function and r(.) is the gamma function. The parameter l is associated with the length scale, while *v* controls the smoothness of the resulting function. Utilizing these specific parameters, we enable a single restart of the optimizer to fine-tune the kernel’s hyperparameters, aiming to improve the model fit. Additionally, we normalize the target values to have zero mean and unit variance, a standard practice to enhance numerical stability and the efficiency of the optimization process. We implement this approach using the *scikit-learn* library [80], which enables us to develop a nonlinear model that predicts the WPI based on DBS settings and quantifies the uncertainty of these predictions. The approach is particularly well-suited for our objective of generating personalized maps of DBS settings to WPI, offering insights into the optimal stimulation parameters for enhancing walking performance. This process continues by incorporating additional visits and experimenting with new settings. Ultimately, we validate the model by evaluating these optimized settings and comparing them to their clinical configurations.

To assess the relationships between the ranked WPI values and feedback rankings from both patients and physical therapists, we employed Spearman’s rank correlation coefficient (*p*). This non-parametric measure of correlation evaluates the strength and direction of association between two ranked variables, making it suitable for ordinal data.

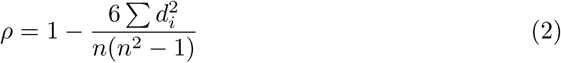

where d_i_ is the difference between the ranks of corresponding variables and n is the number of observations. We selected this approach for its robustness to non-linear relationships and its ability to handle ordinal data, which is appropriate given the ranked nature of both our WPI values and subjective feedback rankings. Within each visit, WPI values were ranked such that the highest WPI value received a rank of 1, with the ranking continuing in descending order. This provided a set of ranked WPI values for each visit. The ranked WPI values and the corresponding feedback rankings from both patients and physical therapists across all subjects were combined into a single dataset for comprehensive analysis.

### Neural data processing and neurophysiological analysis

To synchronize LFP data and ECoG recordings with gait kinematic measurements, we investigated the peak acceleration moments detected by RC+S, Delsys Trigno, and Xsens sensors. Neural data were sampled at 500 Hz and processed through a 1 Hz high-pass filter. We then used MATLAB’s built-in continuous wavelet transform function *(cwt)* for time-frequency analysis. Additionally, we conduct a comprehensive analysis to examine the interaction between cortical and subcortical areas within each hemisphere. This includes coherence between the LFP recordings from the pallidum and ECoG recordings from the motor cortex within each brain hemisphere. To achieve precise wavelet coherence analysis, we utilize MATLAB’s built-in function *(wcoherence)*.

Given the susceptibility of neural signals to various artifacts, we closely examined the time-frequency plots to detect gamma-band artifacts, identifying segments with unusually high-frequency components. We applied a Butterworth high-pass filter (cutoff: 4 Hz, order: 4) using the *Fieldtrip* toolbox to eliminate low-frequency noise. Next, we performed bandpass filtering in the gamma range (75-150 Hz) and calculated *z-scores*, marking data points exceeding an 8 *z-score* threshold as artifacts. We incorporated a 0.2-second blanking buffer around each detected artifact to mitigate spectral artifacts. The identified artifacts were replaced with NaNs, ensuring cleaner data for subsequent analyses. We also addressed potential EKG-induced artifacts by implementing an adaptive EKG artifact removal technique, ensuring accurate identification of EKG artifact instances [81]. Significant EKG artifacts were not prevalent in the conventional sensing channels within this cohort. However, when EKG-like artifacts were detected in stimulation contacts, we carefully analyzed these instances and cross-referenced them with other channels to confirm the absence of EKG artifacts in the LFP recordings. Additionally, we mitigated the possibility of data packet loss in the RC+S devices by identifying such events through low-frequency analysis, thereby excluding the affected gait events from further analyses.

We utilized the *z-score* method for each frequency to normalize the signal power and coherence values within each visit. Next, we extracted single gait trials from the clean, normalized time-frequency representations of neural recordings, and sorted the gait cycles by their stride times. To ensure our conclusions are unaffected by stimulation-induced sub-harmonics artifacts, we focused on the 1-30 Hz canonical frequency bands: delta (2-4 Hz), theta (4-8 Hz), alpha (8-12 Hz), and beta (12-30 Hz). Within these frequency bands, we compute the average levels of signal power during phases of gait cycles across all trials (i.e., exemplified in Figure 4). Furthermore, to assess the impact of variations in DBS settings on the neural signal power across different gait phases, we determined the average levels of signal power for each canonical frequency band during each gait phase. Different phases include two double limb support periods and two swing phases (i.e., right toe off to right heel strike for right leg swing and left toe off to left heel strike as the left leg swing).

### Correlation Between Neural Oscillations and Improved Walking Performance

To determine the neurophysiological basis underlying changes in gait metrics, we analyzed fluctuations in pallidal LFPs, cortical activity, and pallidal-cortical coherence dynamics throughout each gait cycle as participants walked back and forth during the walking performance assessment. We then used linear mixed-effects models to evaluate neural biomarkers of walking performance. This approach included fixed effects for neural features, which represented the influence of neural oscillations on walking performance, and random effects for each individual to account for inter-subject variability. Given the exploratory nature of our analysis, we assessed correlations between individual neural features and walking performance independently, rather than constructing a comprehensive model incorporating all potential features simultaneously. Our aim was to identify consistent neural biomarkers across all participants that correlated with walking performance.

Neural features analyzed included the average levels of spectral power in the canonical frequency bands and coherence between cortical and subcortical regions. These analyses were performed during different phases of the gait cycle. Additionally, we conducted both group-level and person-specific analyses—group-level analyses to identify shared neural markers across the cohort and individual-specific analyses to capture unique characteristics that contributed to improved walking performance under gait-optimized DBS settings. This dual approach ensured a comprehensive understanding of the common and individual neural dynamics linked to gait enhancement.

## 7 Data availability

Data from this study can be made available upon reasonable request, provided that patient confidentiality is maintained and disclosure standards are met.

## 8 Acknowledgements

We appreciate the participants for their involvement in this study. This study is supported in part by NIH grant R01NS130183 (R01), MJFF grant MJFF-010435 (MJFF), and the Burroughs Wellcome Fund Career Award for Medical Scientists. All funding was acquired by D.D.W.

## 9 Ethics declarations

D.D.W. consults for Medtronic, Boston Scientific Corp, and Iota. P.A.S. receives support from Medtronic and Boston Scientific for fellowship education. S.L. consults for Iota Biosciences. H.F., K.H.L., J.E.B., K.N.P., J.P.B., J.H.M., T.A.W., N.B.G., M.S. declare no competing interests.

### 9.1 Ethical approval

This study was reviewed and approved by the Institutional Review Board (IRB) of the University of California, San Francisco (UCSF), under the approval number 20-32847. The UCSF IRB conducted a full board review and granted approval, confirming that the study met all ethical requirements.

## Notes

### Clinical Trial

NCT03582891

### Author Declarations

Participants were enrolled in a clinical trial (ClinicalTrials.gov ID: NCT-03582891) at the University of California San Francisco (UCSF), which was approved by the UCSF Institutional Review Board (IRB) under the approval number 20-32847.

