## Supplementary materials for "Modeling and Optimizing Deep Brain Stimulation to Enhance Gait in Parkinson’s Disease: Personalized Treatment with Neurophysiological Insights"

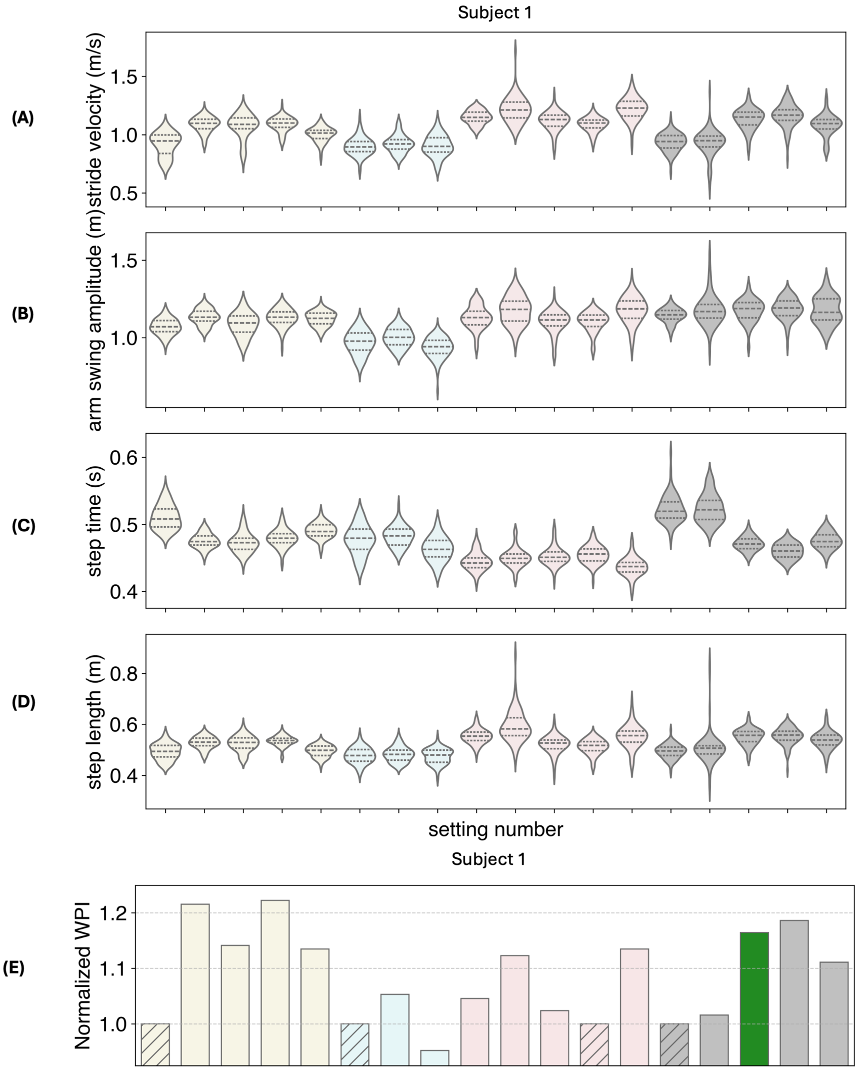


| Visit | Visit 1 | | | | | Visit 2 | | | Visit 3 | | | | | Visit 4 (test & validation) | | | | |
|---|---|---|---|---|---|---|---|---|---|---|---|---|---|---|---|---|---|---|
| Setting | 1 | 2 | 3 | 4 | 5 | 6 | 7 | 8 | 9 | 10 | 11 | 12 | 13 | 14 | 15 | 16 | 17 | 18 |
| Frequency | 150 | 150 | 60 | 60 | 190 | 150 | 190 | 190 | 150 | 60 | 60 | 150 | 190 | 150 | 190 | 60 | 125 | 190 |
| Amplitude | 5.5 | 4.1 | 4.1 | 5.5 | 5.5 | 5.5 | 5.5 | 4.1 | 4.1 | 4.1 | 5.5 | 5.5 | 4.1 | 5.5 | 5.5 | 5.1 | 4.1 | 5.5 |
| Pulse width | 90 | 90 | 90 | 90 | 90 | 90 | 70 | 70 | 90 | 90 | 90 | 90 | 70 | 90 | 70 | 90 | 80 | 80 |

**Fig. S1 Impact of DBS setting adjustments on gait kinematics and walking performance (subject 1)**. Panels **(A)**-**(D)** illustrate the variations in gait kinematics as a response to different DBS settings across three visits, measured by **(A)** stride velocity, **(B)** arm swing amplitude, **(C)** step length variability, and **(D)** step time variability. Panel **(E)** displays the corresponding changes in the walking performance index (WPI). Columns with hatched patterns represent the clinically optimized DBS settings tested at each visit. In Visit 4, the green-colored bars correspond to the DBS settings predicted by the Gaussian Process Regression (GPR) model to enhance walking performance. The detailed stimulation parameters for each setting are summarized in the table at the bottom, with each color representing data from a specific visit.

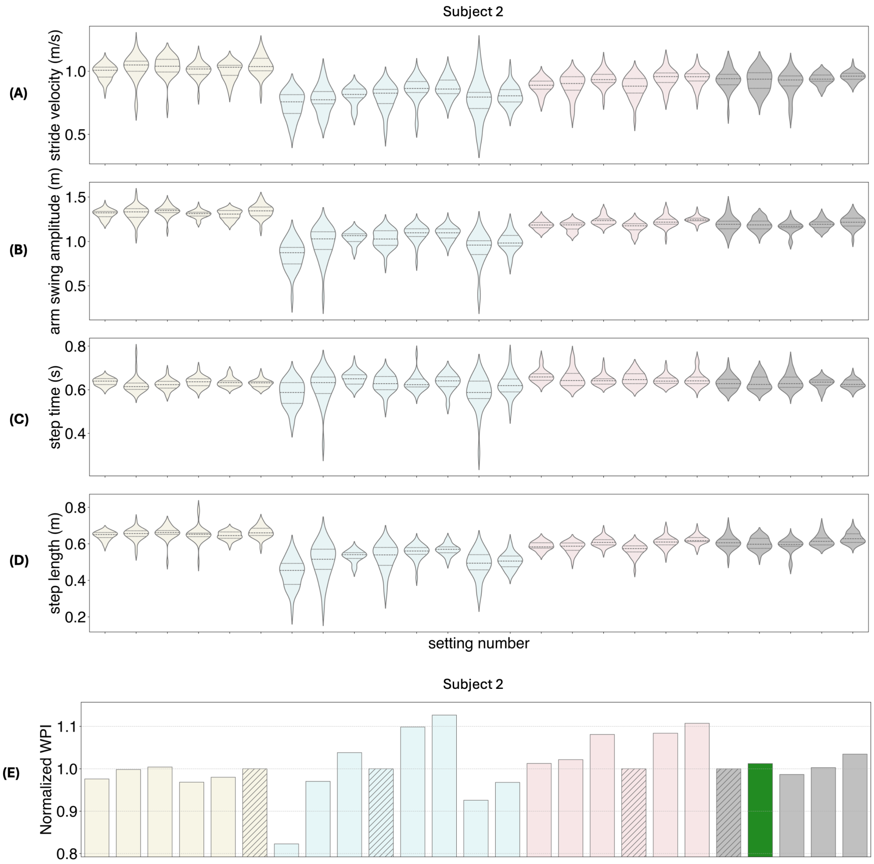


| Visit | Visit 1 | | | | | | Visit 2 | | | | | | | | Visit 3 | | | | | | Visit 4 (test & validation) | | | | |
|---|---|---|---|---|---|---|---|---|---|---|---|---|---|---|---|---|---|---|---|---|---|---|---|---|---|
| Setting | 1 | 2 | 3 | 4 | 5 | 6 | 7 | 8 | 9 | 10 | 11 | 12 | 13 | 14 | 15 | 16 | 17 | 18 | 19 | 20 | 21 | 22 | 23 | 24 | 25 |
| Frequency | 130 | 60 | 60 | 180 | 180 | 130 | 130 | 60 | 60 | 130 | 60 | 60 | 130 | 130 | 130 | 60 | 60 | 130 | 60 | 180 | 130 | 60 | 60 | 180 | 180 |
| Amplitude | 2.8 | 2.8 | 4.0 | 4.0 | 2.8 | 4.0 | 4.0 | 4.0 | 4.0 | 4.0 | 5.0 | 5.0 | 5.0 | 5.0 | 4.0 | 4.0 | 2.8 | 4.0 | 2.8 | 4.0 | 4.0 | 4.0 | 4.8 | 4.0 | 2.8 |
| Pulse width | 60 | 60 | 60 | 60 | 60 | 60 | 70 | 70 | 60 | 60 | 60 | 70 | 70 | 60 | 70 | 70 | 70 | 60 | 60 | 60 | 60 | 60 | 70 | 60 | 60 |

**Fig. S2 Impact of DBS setting adjustments on gait kinematics and walking performance (subject 2)**. Panels **(A)**-**(D)** illustrate the variations in gait kinematics as a response to different DBS settings across three visits, measured by **(A)** stride velocity, **(B)** arm swing amplitude, **(C)** step length variability, and **(D)** step time variability. Panel **(E)** displays the corresponding changes in the walking performance index (WPI). Columns with hatched patterns represent the clinically optimized DBS settings tested at each visit. In Visit 4, the green-colored bars correspond to the DBS settings predicted by the Gaussian Process Regression (GPR) model to enhance walking performance. The detailed stimulation parameters for each setting are summarized in the table at the bottom, with each color representing data from a specific visit.

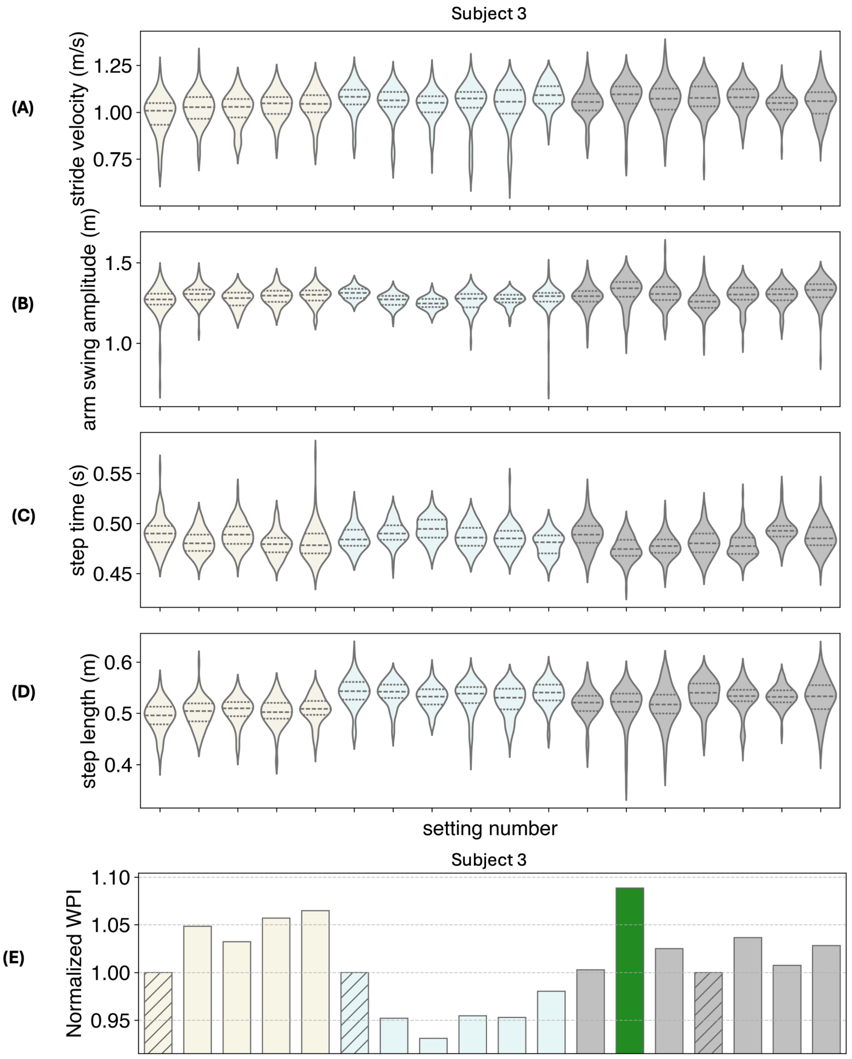


| Visit | Visit 1 | | | | | Visit 2 | | | | | | Visit 3 (test & validation) | | | | | | |
|---|---|---|---|---|---|---|---|---|---|---|---|---|---|---|---|---|---|---|
| Setting | 1 | 2 | 3 | 4 | 5 | 6 | 7 | 8 | 9 | 10 | 11 | 12 | 13 | 14 | 15 | 16 | 17 | 18 |
| Frequency | 130 | 60 | 60 | 130 | 180 | 130 | 60 | 60 | 130 | 180 | 180 | 60 | 180 | 145 | 145 | 190 | 60 | 145 |
| Amplitude | 4.7 | 4.7 | 3.5 | 3.5 | 3.5 | 4.9 | 4.9 | 3.6 | 3.6 | 3.6 | 4.9 | 3.9 | 4.2 | 3.0 | 3.9 | 3.5 | 3.9 | 4.9 |
| Pulse width | 60 | 60 | 60 | 60 | 60 | 60 | 60 | 80 | 80 | 80 | 60 | 60 | 80 | 60 | 60 | 80 | 80 | 80 |

**Fig. S3 Impact of DBS setting adjustments on gait kinematics and walking performance (subject 3)**. Panels **(A)**-**(D)** illustrate the variations in gait kinematics as a response to different DBS settings across three visits, measured by **(A)** stride velocity, **(B)** arm swing amplitude, **(C)** step length variability, and **(D)** step time variability. Panel **(E)** displays the corresponding changes in the walking performance index (WPI). Columns with hatched patterns represent the clinically optimized DBS settings tested at each visit. In Visit 3, the green-colored bars correspond to the DBS settings predicted by the Gaussian Process Regression (GPR) model to enhance walking performance. The detailed stimulation parameters for each setting are summarized in the table at the bottom, with each color representing data from a specific visit.

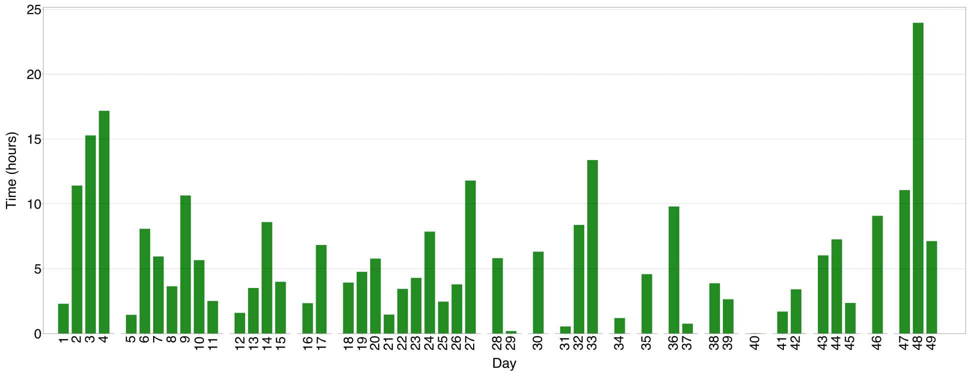


**Fig. S4 Time spent on gait-optimized setting (subject 1).** The bar chart illustrates the total time intervals measured across consecutive days. Sequential day numbers are displayed on the x-axis for days with recorded data. Gaps in the data are visually represented by open spaces on the x-axis, indicating periods where no changes to the DBS settings were made. The y-axis represents the total time interval (in hours) for each day.

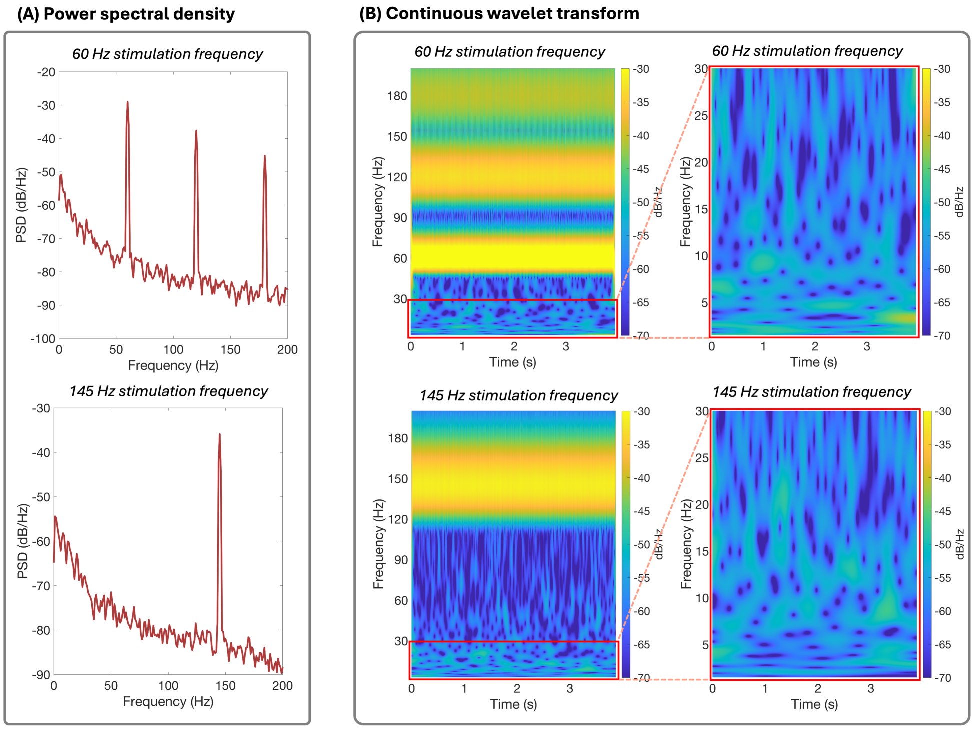


**Fig. S5 Impacts of stimulation-induced artifacts in time-frequency analysis.** Panel **(A)** illustrates the power spectral density of the LFP signal from the GP under two stimulation conditions. The top sub-panel shows the results at a stimulation frequency of 60 Hz, while the bottom sub-panel shows results at 145 Hz. Panel **(B)** presents the continuous wavelet transformation of the GP signal, with the top and bottom sub-panels corresponding to the 60 Hz and 145 Hz stimulation frequencies, respectively. The right sub-panels in panel **(B)** display a zoomed-in view of the frequency range between 0 and 30 Hz, highlighting the focus of the analysis on avoiding stimulation-induced artifacts in this lower frequency band.

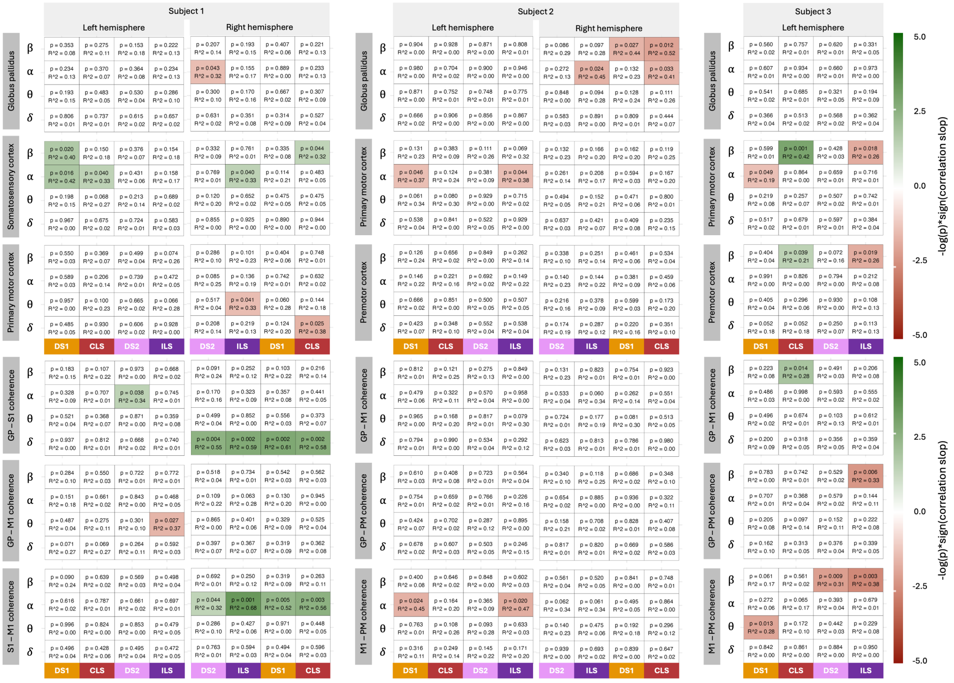


**Fig. S6 Comparison of neural features correlating with walking performance across subjects.** Correlations were assessed using linear regression between neural features (average levels of neural signals from various gait phases across canonical frequency bands) and the Walking Performance Index (WPI). The top three rows show the correlations between neural signals from the Globus Pallidus and motor cortices, while the bottom three rows depict the correlations between coherence levels and WPI. Each tile represents the statistical significance (p-value) and goodness-of-fit ($R^2$) of the linear regression models for different frequency bands (y-axis) and gait phases (x-axis). Canonical frequency bands include $\delta$ (2-4 Hz), $\theta$ (4-8 Hz), $\alpha$ (8-12 Hz), and $\beta$ (12-30 Hz). Gait phases are represented as DLS1 (double limb support 1) from LHS to RTO, CLS (contralateral leg swing) from RTO to RHS, DLS2 (double limb support 2) from RHS to LTO, and ILS (ipsilateral leg swing) from LTO to LHS for signals from the left hemisphere (all subjects). For the right hemispheres (subjects 2 and 3), gait phases are represented as DLS2 (double limb support 2) from LHS to RTO, ILS (ipsilateral leg swing) from RTO to RHS, DLS1 (double limb support 1) from RHS to LTO, and CLS (contralateral leg swing) from LTO to LHS. The color of the tiles indicates the value of $-log_{10}$(p-value)*$sign$(slope), with deeper red representing a negative association and deeper green representing a positive association. We show $-log_{10}$(p-value), where values below -1.3 and above 1.3 indicate significance at the $p < 0.05$ level. Values for non-significant features ($p > 0.05$) are set to zero, with white tiles indicating non-significant features.
